# Ostavimir is ineffective against COVID-19: in silico assessment, in vitro and retrospective study

**DOI:** 10.1101/2020.05.15.20102392

**Authors:** Qi Tan, Yang Jin

**Affiliations:** Department of Respiratory and Critical Care Medicine, NHC Key Laboratory of Pulmonary Disease; Wuhan Union Hospital

## Abstract

As a neuraminidase inhibitor, oseltamivir has effectively combated the pandemic influenza A and B, so it is a first-line commonly used antiviral drug, especially in primary hospitals. At the same time, oseltamivir, as an over-the-counter drug, is also a popular antiviral drug. As healthcare workers fighting against coronavirus disease 2019 (COVID-19), we have found that many patients experiencing discomfort or considered to be infected with a virus take oseltamivir. From severe acute respiratory syndrome coronavirus (SARS-CoV) in 2003 to middle east respiratory syndrome coronavirus (MERS-CoV) in 2012, and now the current COVID-19 epidemic, there is not plenty of evidence showing that oseltamivir is effective against coronavirus. Still, there is also no sufficient evidence to refute its ineffectiveness. We cannot predict whether there will be a pandemic of respiratory coronavirus in the future, so we hope to initiate such research and preliminarily explore whether oseltamivir is effective for COVID-19, which can better guide healthcare workers in the selection of appropriate antiviral drugs in the face of coronavirus epidemics. If oseltamivir is effective, then a wide promotion of its application often can achieve a double effect with half the effort. If it is not effective, then considering the side effects of oseltamivir, it is not necessary to use unreasonable drugs that will not slow the progression of the disease but can cause adverse reactions. We found that oseltamivir isn’t suitable for fighting against COVID-19 through the method of computer aided drug design and in vitro study and retrospective case study. Meanwhile it was high-occurrence seasons for the influenza, COVID-19 should be highly suspected in patients who didn’t benefit from oseltamivir. We hope that the result of our study could be shared with the frontline physicians in fighting against COVID-19.

---

As a neuraminidase inhibitor, oseltamivir was approved by the Food and Drug Admission (FDA) in 1999^1^. Since then, it has played an essential role in treating against influenza A and influenza B and is becoming more widespread. The atypical pneumonia caused by severe acute respiratory syndrome coronavirus (SARS-CoV) that broke out in Guangzhou, China, in 2003, linked oseltamivir to coronavirus. Zhang et al. found that the active site of the Spike (S) 1 Protein of SARS is similar to that of neuraminidase, suggesting that neuraminidase inhibitors may be useful to treat SARS-CoV^2^. However, despite this similarity, no clinical data suggest that oseltamivir is effective in treating SARS-CoV. With the epidemic of coronavirus disease 2019 (COVID-19) caused by SARS-CoV-2, oseltamivir has once again become a hot topic. A study from Thailand reported that a 71-year-old patient with severe COVID-19 on January 29, 2020 underwent a 48-hour treatment with lopinavir/ritonavir combined with oseltamivir, which improved the patient’s condition, and the throat swab test became negative^3^. Although a single case report does not prove the effectiveness of oseltamivir for COVID-19, it once again linked oseltamivir to the treatment of a coronavirus-induced disease. As frontline healthcare workers fighting against COVID-19, we were interested in the treatment regimen of this case and found most patients with COVID-19 who were symptomatic have used oseltamivir. We believed that it is of practical significance to further study the effect of oseltamivir on COVID-19. Because of its significant effectiveness on influenza and being sold over the counter, oseltamivir is not only stocked in common households but also a common antiviral drug in primary hospitals. Therefore, if oseltamivir is effective for COVID-19 the treatment could be family-oriented. If, instead, the treatment effectiveness is not significant, the use of oseltamivir should be stopped, which will avoid delaying other treatments and multiple oseltamivir adverse reactions, such as nausea, vomiting, epilepsy, elevated liver enzymes, and arrhythmias^4,5^. Therefore, the study of the antiviral effect of oseltamivir against SARS-CoV-2 could have a positive effect on the treatment of COVID-19.

SARS-CoV-2 belongs to the β-genus of the coronavirus family and is the seventh coronavirus found to infect humans. Therefore, it has the structural features of coronaviruses, such as structural proteins, including S protein and Nucleoprotein (NC), and also non-structural proteins, including the main key enzymes 3C-like protease (3CLpro), papain-like protease (PLpro), and RNA-directed RNA polymerase (Pol/RdRp). These main structural proteins and key enzymes of SARS-CoV-2 are important targets for inhibiting SARS-CoV-2 infection^6–9^. Since the SARS outbreak in 2003, the S protein, NC, 3CLpro, PLpro, and Pol/RdRp of the coronavirus known to infect humans have been extensively studied due to their potential as therapeutic targets^10–15^.

Oseltamivir is a competitive inhibitor of neuraminidase designed by computer-assisted technology. In this context, the effective binding of oseltamivir to the active site of key proteins of SARS-CoV-2 is particularly important. Therefore, here we: 1. analyzed whether S protein, NC protein, 3CLpro, PLpro, and Pol/RdR have active centers similar to neuraminidase; 2. explored whether oseltamivir can form effective docking with key protein active centers of the virus using molecular simulation; 3. accessed the antiviral effect of oseltamivir against SARS-CoV-2 in vitro; 4. used case review to analyze whether oseltamivir is a factor to improve the conditions of patients with COVID-19.

## The obtainment of the viral protein structure

The structures of the S protein (PDB ID: 6VSB) and 3CL pro (PDB ID: 6LU7) of SARS-CoV-2 were obtained from the RCSB Protein Data Bank (http://www.rcsb.org). Then the structures of NC, PLPro, and Pol/RdRp were built through the method of homology modeling. We first downloaded the protein sequences of NC (YP_009724397.2), nsp3 (YP_009725299.1), and Pol/RdRp (YP_009725307.1) from the GenePept protein sequence database and used Swiss-model (www.swissmodel.org) program for homology modeling. The models were constructed successfully and shown in Fig1A-C. Among them, the NC of SARS-CoV-2 contains 419 amino acid residues. The search of a homology template using the Swissmodel found that the 35–174 residues of NC have an identity of 92.37% with the N-terminal RNA-binding domain (NRBD) of NC from SARS-CoV(PDB ID: 1ssk), which is the active region for the binding of NC of SARS-CoV with RNA. Inhibiting NRBD activity can affect coronavirus RNA transcription and replication, and virion assembly^16^. Thus, it is one of the targets for inhibiting coronaviruses. Therefore, NRBD is the main activity center of NC, and this model can be used for the active site of NC from SARS-CoV-2 for further research.

**Figure 1.**
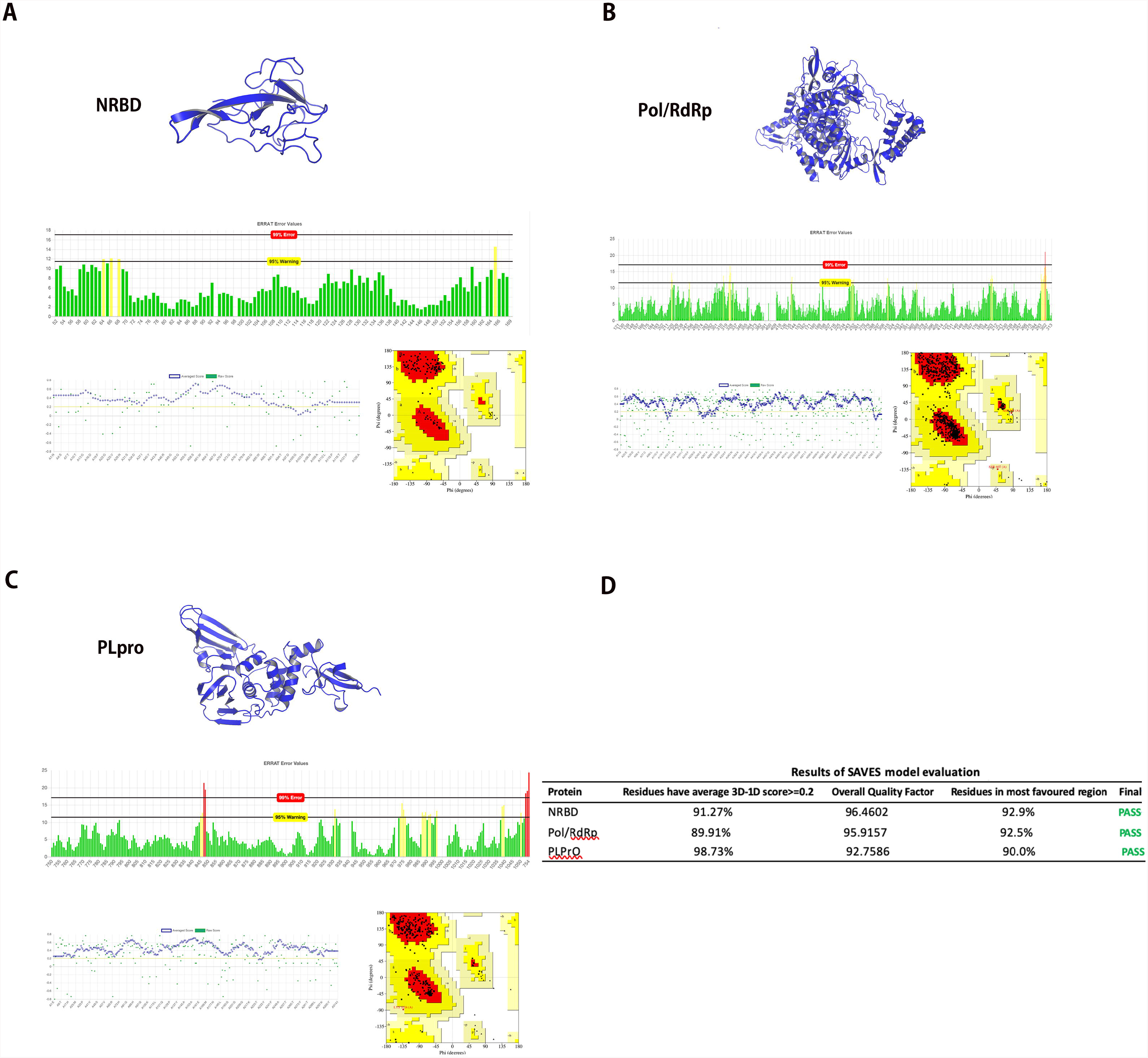
A. The model of NRBD from SARS-CoV-2 with the evaluation of EBRAT, Verify3D, and Ramachandran plot; B. The model of Pol/RdRp of SARS-CoV-2 with the evaluation of EBRAT, Verify3D, and Ramachandran plot; C. The model of PLpro from SARS-CoV-2 with the evaluation of EBRAT, Verify3D, and Ramachandran plot; D Result of SAVES model evaluation. NRBD: N-terminal RNA-binding domain; Pol/RdRp: RNA-directed RNA; polymerase; PLpro: papain-like protease;

## Evaluation of virus models

Subsequently, we used the SAVES v5.0 program (servicesn.mbi.ucla.edu/SAVES/) to carry out a model evaluation, and the evaluation criteria for a successful model were as follows: 1. in ERRAT score result, the Overall Quality Factor was >85; 2. in the Verify3D assessment results, > 80% of the residues have average 3D-1D score ≥ 0.2; 3. in the Procheck assessment result, ≥90% residues were in the most favored region. The results are shown in Fig 1D, suggesting all three models passed the evaluation. Among them, the ERRAT score was based on whether the error value of the alignment of the non-bonding interaction among the atoms in the structure and the high-resolution crystal structure was within a reasonable confidence interval. If the percentage of the reasonable confidence interval is higher, the quality is higher. Almost all residues of NRBD, Pol/RdRp, and PLpro were within reasonable confidence intervals, and the final scores were 96.4602, 95.9157, and 92.7586, respectively (Fig1A-C). Verify3D was used to analyze the compatibility of the 3D structure and the primary sequence in the model. A 3D-1D score ≥ 0.2 of a residue is reasonable. The higher the ratio of reasonable residues, the higher the quality of the model. The majority of residues of NRBD, Pol/RdRp, and PLpro belonged to reasonable residues (green), and there were 91.27%, 89.91%, and 98.73% reasonable residues in NRBD, Pol/RdRp, and PLpro respectively (Fig1A-C). The results of Procheck were mainly shown by Ramachandran plots. As can be seen in the Ramachandran plot, there were A, B, L, a, b, l, p, ∼a, ∼b, ∼p, and ∼l labeled areas. In Porcheck, A, B, and L are classified as reasonable areas; a, b, l are relatively reasonable regions, whereas ∼a, ∼b, ∼p, and ∼l are acceptable regions. The distributions of amino acid residues in the structure were observed. The large proportion of amino acids in reasonable regions proves that the model is of high quality. It can be seen that most of the residues of NRBD, Pol/RdRp, and PLpro were in A, B, and L, and the reasonable region residues accounted for 92.9%, 92.5%, and 90.0%, respectively (Fig1A-C). The above results suggested that the three modeled structures passed the evaluation, were of good quality, and could be used for further research.

## Alignment of the structure of viral proteins and influenza A neuraminidase

To explore whether oseltamivir is effective for SARS-CoV-2, we first analyzed whether there was structural similarity between the structure of viral protein and influenza A neuraminidase (PDB ID: 3Ti6), especially whether there was a similarity in the active center. TM-align program (https://zhanglab.ccmb.med.umich.edu/TM-align/) was used for structural alignment^17^. The alignment of the structure of influenza B neuraminidase (PDB ID: 4CPY) with 3Ti6 was selected as a positive control. TM-score was used to analyze the structural similarity; 0.0 < TM-score < 0.30 was defined as “random structural similarity”, and 0.5 < TM-score < 1.00 was defined as “in about the same fold”. The active site of influenza A and B neuraminidase were determined based on the location of ligand in crystal structure (Fig. 2A)^18,19^. By comparing 4CPY with 3Ti6, the TM-score of the two was found to be 0.94460, which proved that the two were structurally homologous. Subsequently, we aligned the structures of S protein, NRBD, 3CL pro, PLpro, and Pol/RdRp with 3Ti6; the TM-scores were 0.30077, 0.19254, 0.28766, 0.30666, and 0.34047, respectively (Fig. 2G). The results suggested that there was only a structural similarity between the protein structure of the above SARS-CoV-2 and 3Ti6. The active center of the S protein was mainly the RBD domain (Fig. 2B green region) and S2 subunit HR1 domain (Fig. 2B blue region), which were also two important targets for studying the inhibition of S protein^20,21^. We found that the similar structure of 3Ti6 and S protein is located in the N-terminal domain (NTD) region of S protein. As the active center of NC, we found that NRBD did not match the active center of 3Ti6 (Fig2C). The active center of 3CLpro was determined based on that it is homologous to the 3CLpro from SARS-CoV. Previous studies have confirmed the active center of 3CLpro of SARS-CoV. Thus, this region was used as the active center of 3CLpro of SARS-CoV-2^12^. By alignment, we found that the active centers of the two proteins were similar (Fig. 2D). Subsequently, we also determined the active center of the PLpro and Pol/RdRp based on the homology of the proteins of SARS-CoV-2 and those of SARS-CoV. And we found that the active center of PLpro did not match the active center from 3Ti6 through the alignment of structure (Fig. 2E)^22^. In studies focused on the Pol/RdRp of SARS-CoV, it has found that Thr680, Asn691, and Asp623 can bind nucleotides and promote viral gene transcription, and Val557 also plays an important role in the replication of the virus^19^. Thus, the region where the above residues are located was determined as the active center of SARS-CoV-2 (Fig. 2F). It was found that the similar structure between Pol/RdRp and neuraminidase was not located in the active center. The above results suggested that there was a theoretical possibility of effective binding of oseltamivir with the active center of 3CLpro because it was found to be similar to the active center from the neuraminidase.

**Figure 2.**
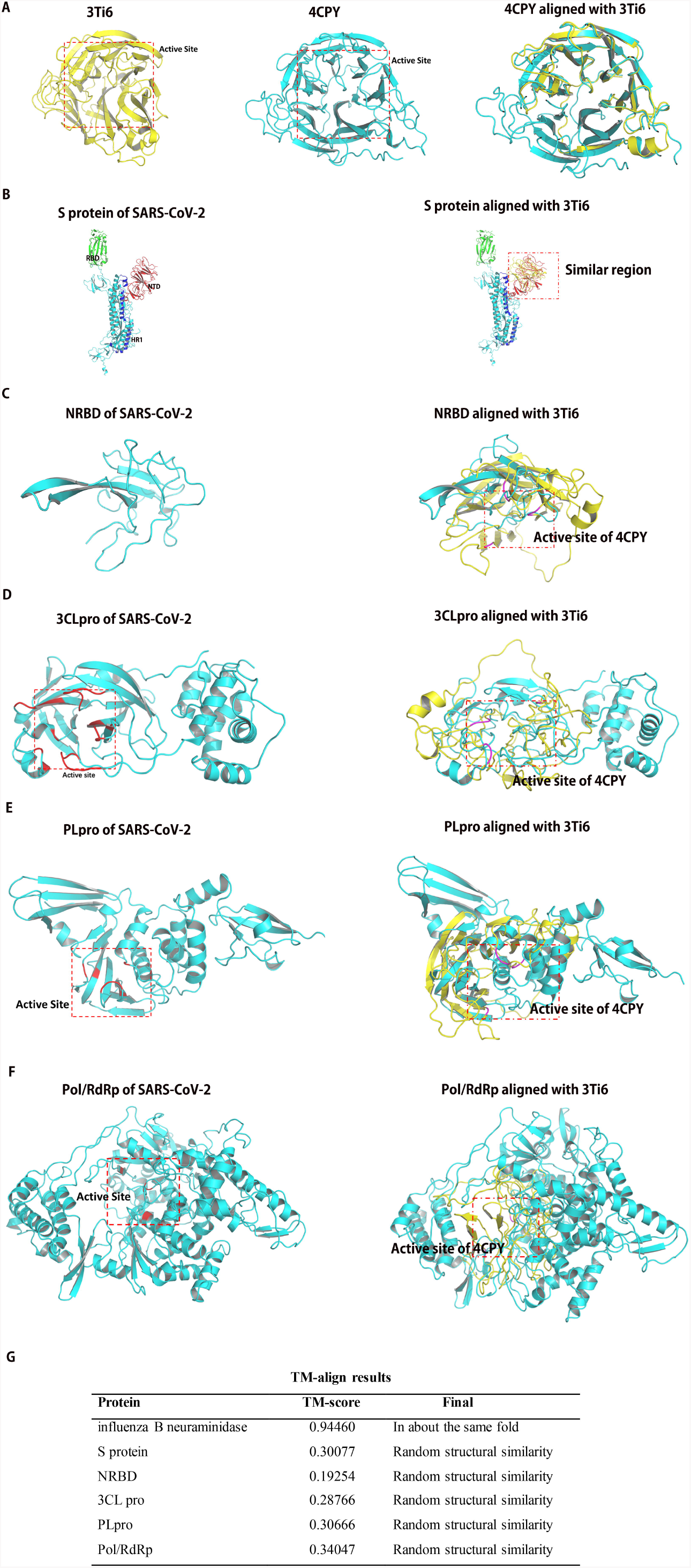
A: Alignment of the structure of influenza A neuraminidase and influenza B neuraminidase. B: Alignment of the structure of S protein and influenza A neuraminidase; C: Alignment of the structure of NRBD and influenza A neuraminidase; D: Alignment of the structure of 3CLpro and influenza A neuraminidase; E: Alignment of the structure of PLpro and influenza A neuraminidase; F: Alignment of the structure of Pol/RdRp and influenza A neuraminidase; G: Results of TM-align. NRBD: N-terminal RNA-binding domain; Pol/RdRp: RNA-directed RNA polymeras; PLpro: papain-like protease; 3CLpro:3C-like protease; S protein: Spike Protein;

## Molecular Docking Studies for predicting the antiviral effect of oseltamivir

To pursue our hypothesis, we used molecular docking to analyze whether oseltamivir carboxylic acid could stably bind to the active center of the viral protein structure. We used ChemBioDraw Ultra 17.0 software to draw the structure of oseltamivir carboxylic acid and N3 inhibitor, then converted them into three-dimensional (3D) structure using ChemBio3D Ultra 17.0 software. Then the structure of agents was preprocessing by MMFF94 force field. AutoDock Tools 1.5.6 software was used to remove water molecules and ligands in the protein, and then add hydrogen atoms, combine non-polar hydrogen atoms, add charges, and converted protein and drugs into the format of PDBQT. The GridBox module was used to generate a 3D ensemble around the active site, and the grid points in the ensemble were calculated for energy score evaluation. AutoDock Vina was used for molecular docking^23^. Exhaustiveness was set at 20 and other parameters were used default value. Finally, we selected the conformation with the highest docking score to analyze the result using Free Maestro 11.9. The docking conformations were described in Fig3A-E. The results showed that the docking energies of oseltamivir carboxylic acid with NRBD, 3CLpro, PLpro, and Pol/RdRp were −4.7 kcal/mol, −7.0 kcal/mol, −5.8 kcal/mol, and −5.4 kcal/mol, respectively (Fig3F). The lower the binding energy, the more suitable the binding of the inhibitor molecule to the receptor. In terms of binding energy, oseltamivir carboxylic acid was more favorable to bind to the active site of 3CLpro effectively. However, we concluded that the ability of oseltamivir to inhibit 3CLpro is not strong compared with N3 inhibitor, which exhibited strong inhibition for 3CLpro of SARS-CoV-2 in vitro and its docking energy was −9.5 kcal/mol^24^, which was lower than that of oseltamivir carboxylic acid.

**Figure 3.**
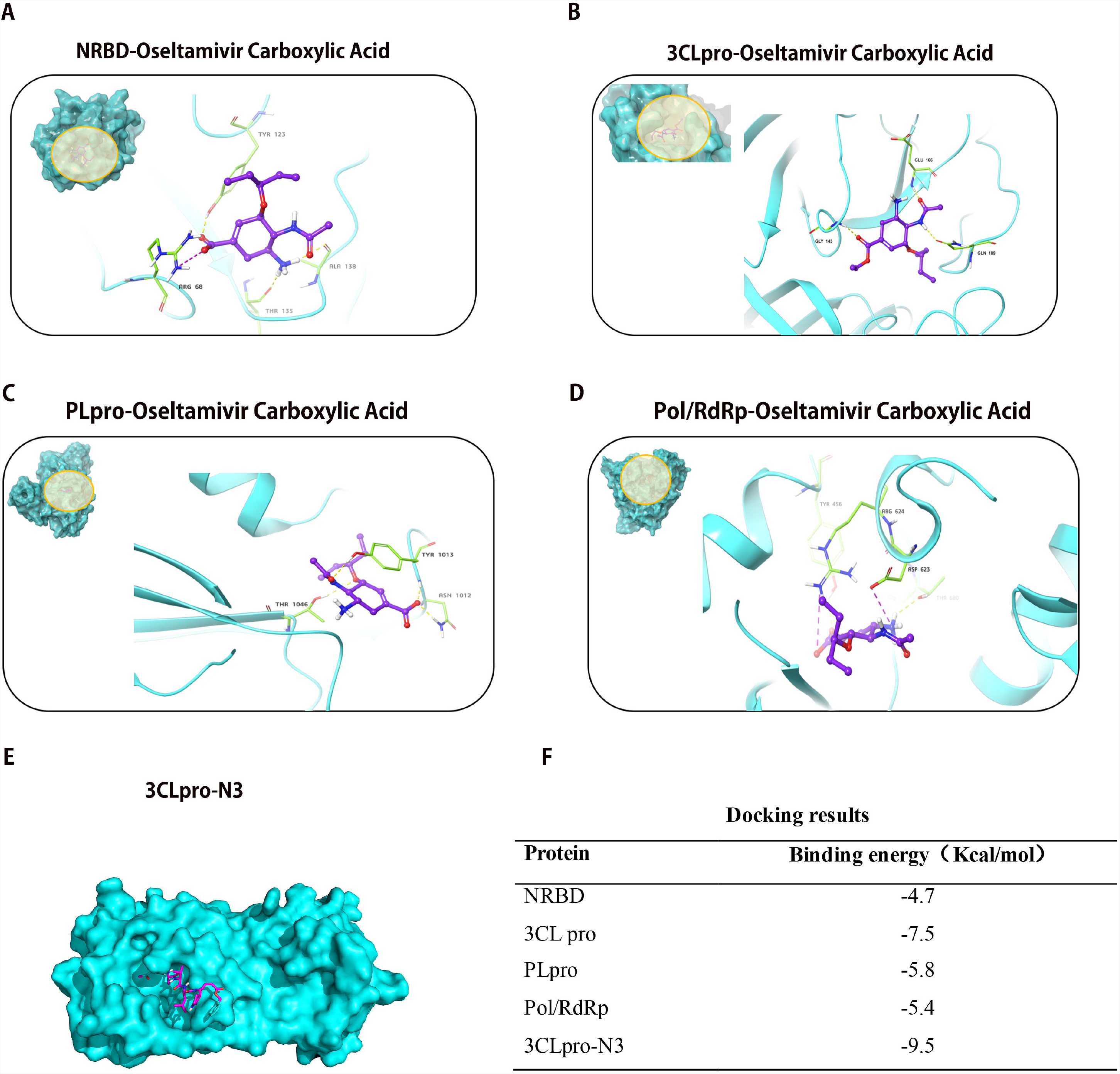
A: Docking conformation of NRBD and oseltamivir carboxylic acid; B: Docking conformation of 3CLpro and oseltamivir carboxylic acid; C: Docking conformation of PLpro and oseltamivir carboxylic acid; D: Docking conformation of Pol/RdRp and oseltamivir carboxylic acid; E: Docking conformation of 3CLpro and N3 inhibitor; F: Results of Docking. NRBD: N-terminal RNA-binding domain; Pol/RdRp: RNA-directed RNA polymerase; PLpro: papain-like protease; 3CLpro: 3C-like protease; S protein: Spike Protein;

## In vitro study for accessing the antiviral effect of oseltamivir

To further verify our hypothesis, we evaluated the antiviral efficiency of oseltamivir against SARS-CoV-2 in vitro as described in the previous work^25^. Briefly, Vero E6 cells infected with the SARS-CoV-2 were cultured with the different doses of the oseltamivir carboxylic acid. Cell culture supernatant was collected after incubated with agent for 24h and viral RNA was extracted. Quantitative real-time RT-PCR was performed to evaluate the quantification of viral copy numbers in the cell supernatant. Fig4A showed that treatment of oseltamivir did not inhibit SARS-CoV-2 replication, with half-inhibitory concentration (IC_50_) value above 100 μM. Chloroquine was selected as a positive control and can inhibit SARS-CoV-2 replication (Fig4B). Therefore, we concluded that oseltamivir is ineffective against SARS-CoV-2.

**Figure 4.**
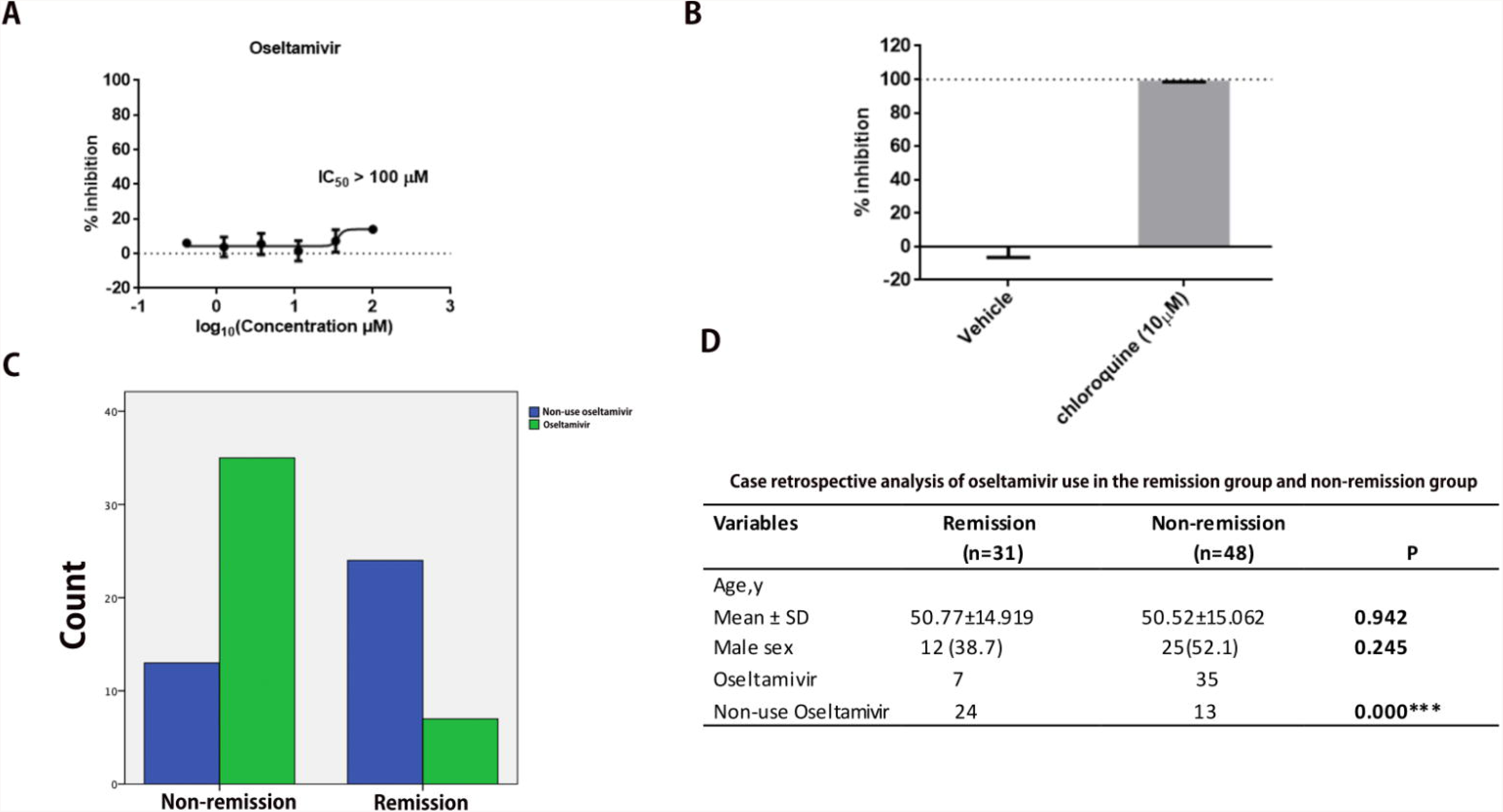
A. The antiviral effect of oseltamivir against SARS-CoV-2 in vitro; B. The antiviral effect of chloroquine against SARS-CoV-2 in vitro; IC50: Half-inhibitory concentration; C-D: Comparison of the number of patient used oseltamivir in the remission group and those in the non-remission group.

## Evaluating the antiviral effect of oseltamivir against SARS-CoV-2 in case review

To further verify the efficacy of oseltamivir on SARS-CoV-2, we retrospectively analyzed a total of 612 patients from the Second Hospital of Wuhan, Wuhan Union Hospital, and Wuhan Union Hospital West Campus, Hubei Province, which was approved by the institutional ethics board of Tongji Medical College, Huazhong University of Science and Technology, Wuhan, China.The enrollment criteria were: 1. patients were diagnosed with COVID-19 and negative for influenza A and B nucleic acid tests; 2. there were no other underlying diseases, such as hypertension, diabetes, cardiovascular and cerebrovascular diseases, malignant diseases, no respiratory tract diseases, such as COPD, asthma, and pulmonary heart disease, and no hypoproteinemia and hypokalemia. The patients’ age and sex were obtained from electronic medical records. We defined the period from the start of antiviral therapy to clinical evaluation as a patients’ treatment period. We defined the patients with relieved symptoms and stable or absorbed lesions in CT during the treatment period as the remission group, and the patients without symptom improvement or no significant changes or progression of the lesions in CT as the progressive group. Whether the history of oseltamivir use influenced the progression of the disease was statistically evaluated. Chi-square test was used to determine differences between group remission and progressive. P-values < 0.05, < 0.01, and < 0.001 were considered significant. Finally, 79 patients were included for analysis. Among them, there were 31 patients in remission, aged 50.77 ± 14.919. There were 48 non-remission patients, and their age was 50.52 ± 15.062. There was no significant difference in age between the two groups. In the remission group, there were 12 males (with a proportion of 38.7%), and in the non-remission group there were 25 males(52.1%); there was no statistical difference in sex between the two groups. We analyzed the use of oseltamivir in the two groups. In the remission group, a total of seven patients used oseltamivir with a proportion of 22.6%. In the non-remission group, a total of 35 patients used oseltamivir, with a proportion of 72.9%. The chi-square value was 19.166, and there was a significant statistical difference between the two groups. The number of patients receiving oseltamivir in the non-remission group was significantly higher than the number in the remission group (P = 0.000) (Fig.4C-D). In combination with clinical practice, we believe that oseltamivir had no inhibitory effect on COVID-19 and could not effectively slow the progression of the disease with early use.

## Conclusion

In this article, we constructed protein models of NRBD, PLpro, and Pol/RdRp of SARS-CoV-2. We used structural alignment to find that the active center of 3CLpro was similar to that of influenza A neuraminidase. Subsequently, we used a molecular docking method to explore whether oseltamivir could effectively bind to the NRBD, 3CLPro, PLpro, and Pol/RdRp of SARS-CoV-2. It was found that oseltamivir carboxylic acid was more favorable to bind to the active site of 3CLpro effectively, but its inhibitory effects was not strong. Finally, we used in vitro study and retrospective case analysis to verify our speculations. We found that oseltamivir is ineffective against SARS-CoV-2 in vitro study and the clinical use of oseltamivir did not improve the patients’ symptoms and signs and did not slow the disease progression. Therefore, we consider that oseltamivir isn’t suitable for the treatment of COVID-19. During the outbreak of novel coronavirus, when oseltamivir is not effective for the patients after they take it, health workers should be highly vigilant about the possibility of COVID-19. At the same time, it should be noted that we are still in the influenza season, and we have also found five cases of influenza A in combined with COVID-19. Therefore, oseltamivir should be combined in the treatment regimen in a timely manner when influenza occurs.

## Data Availability

The data used to support the findings of this study are available from the corresponding author upon request.

